# SARS-CoV-2 vaccine antibody response and breakthrough infection in dialysis

**DOI:** 10.1101/2021.10.12.21264860

**Authors:** Shuchi Anand, Maria E. Montez-Rath, Jialin Han, Pablo Garcia, LinaCel Cadden, Patti Hunsader, Curt Morgan, Russell Kerschmann, Paul Beyer, Mary Dittrich, Geoffrey A Block, Glenn M Chertow, Julie Parsonnet

## Abstract

**Background:** Patients receiving dialysis are a sentinel population for groups at high risk for death and disability from COVID-19. Understanding correlates of protection post-vaccination can inform immunization and mitigation strategies.

**Methods:** Monthly since January 2021, we tested plasma from 4791 patients receiving dialysis for antibodies to the receptor-binding domain (RBD) of SARS-CoV-2 using a high-throughput assay. We qualitatively assessed the proportion without a detectable RBD response and among those with a response, semiquantitative median IgG index values. Using a nested case-control design, we matched each breakthrough case to five controls by age, sex, and vaccination-month to determine whether peak and pre-breakthrough RBD IgG index values were associated with risk for infection post-vaccination.

**Results:** Among 2563 vaccinated patients, the proportion without a detectable RBD response increased from 6.6% [95% CI 5.5-8.1] in 14-30 days post-vaccination to 20.2% [95% CI 17.1-23.8], and median index values declined from 92.7 (95% CI 77.8-107.5) to 3.7 (95% CI 3.1-4.3) after 5 months. Persons with SARS-CoV-2 infection prior-to-vaccination had higher peak index values than persons without prior infection, but values equalized by 5 months (p=0.230). Breakthrough infections occurred in 56 patients, with samples collected a median of 21 days pre-breakthrough. Peak and pre-breakthrough RBD values <23 (equivalent to <506 WHO BAU/mL) were associated with higher odds for breakthrough infection (OR: 3.7 [95% CI 2.0-6.8] and 9.8 [95% CI 2.9-32.8], respectively).

**Conclusions:** The antibody response to SARS-CoV-2 vaccination wanes rapidly, and in persons receiving dialysis, the persisting antibody response is associated with risk for breakthrough infection.

---

Vaccinations are typically preformed on a routine schedule with no post vaccine measurement of immune response. Data linking circulating antibody titers to risk of reinfection are sparse, and among healthy persons, the Advisory Committee on Immunization Practices (ACIP) recommends against checking antibody titers post-vaccination ^1,2^. Post-vaccination circulating antibody titers, however, have been used as correlates of protection in a variety of clinical scenarios^3, 4^.

Among patients on dialysis, there is a precedent for testing response to vaccination in order to inform vaccination schedules^5,6^. Ample data indicate lower rates of seroconversion post hepatitis B and influenza vaccination^7-9^; moreover, the response is shorter in duration compared with that of healthy controls^7^. Thus, patients on dialysis with hepatitis B surface antibody titers below 10 IU/mL two months after the primary vaccination series are revaccinated, or administered a booster if titers (measured annually) fall below 10 IU/mL^6^.

Although a majority of patients receiving dialysis seroconvert post SARS-CoV-2 vaccination, we have previously found that the response was diminished in up to 15% and differed by vaccine type^10^. The duration of circulating antibody levels post-vaccination is unknown. Moreover, evidence from mRNA1273^11^ and ChAd0×1^12^ vaccination randomized control trials indicate a higher risk for post-vaccination (“breakthrough”) infection among persons with lower neutralizing, spike or receptor binding antibody (RBD) titers in the early post-vaccination period. Real world data from BNT162b2-vaccinated Israeli health care workers also described an association between lower peri-infection antibody titers and breakthrough infection^13^. Knowing the strength and duration of antibody response to SARS-CoV-2 vaccination in high-risk groups could help to optimize their immunization schedules and strategies for pre- or post-exposure prophylaxis. In this study we sought to delineate the duration of antibody response to SARS-CoV-2 vaccination among patients receiving dialysis, and to determine whether antibody titers to SARS-CoV-2 could identify patients on dialysis at risk for breakthrough COVID-19 infection.

## Methods

Starting in January 2021, we tested monthly remainder plasma samples from a cohort of persons receiving dialysis at US Renal Care, a dialysis network with over 350 facilities located nationwide. In partnership with Ascend Clinical, a central laboratory processing routine monthly laboratory tests of persons receiving dialysis at several dialysis networks including US Renal Care, we tested these samples for RBD antibody, and ascertained patient characteristics, vaccination status, and COVID-19 diagnoses using electronic health records. The study received ethics approval from Stanford University. Stanford University investigators received anonymized data, and the Institutional Review Board waived requirement for consent.

### Sampling

In the first two weeks of January 2021—prior to widespread COVID19 vaccine roll out—we tested SARS-CoV-2 antibody status of 21,570 patients receiving dialysis^14^. From among the 17,390 seronegative patients, we used systematic sampling with fraction intervals^15^ stratified by age to randomly select 4,346 persons to follow with monthly SARS-CoV-2 serology assays (see **Supplemental Methods** for sample size). We also followed 540 SARS-CoV-2 seropositive patients in whom we knew the date of seroconversion based on our prior work^16^.

### Patient demographics and COVID-19 infection diagnosis

We extracted routinely collected data on age, sex, self-reported race and ethnicity, and diabetes, along with data collected by US Renal Care on date and type of SARS-CoV-2 vaccination. Similarly, US Renal Care instituted a questionnaire-based health screening for all patients; if a patient reported relevant symptoms, dialysis staff requested that the patient be tested. Patients were tested either at the dialysis facility or elsewhere, and results were recorded. In addition to tracking results within each dialysis facility, US Renal Care also tracked whether a patient was hospitalized with COVID-19 or diagnosed with COVID-19 at another healthcare facility. We extracted data on hospitalizations in the 7-day period preceding or 14-day period post COVID-19 infection date.

### Laboratory testing for RBD antibodies

We tested remainder samples using the Siemens’ total RBD Ig assay. The assay is reported by the manufacturer to have 100% sensitivity and 99.8% specificity if performed ≥14 days after a positive reverse transcriptase polymerase chain reaction test (RT-PCR)^17^; it has been validated independently with similar performance characteristics^18,19^. Monthly thereafter we used this assay to test remainder samples from patients in whom antibodies had not been detected in the prior month. Subsequent to a positive total RBD Ig result, the positive sample and all subsequent monthly samples were tested using a semiquantitative Siemens RBD IgG assay. The Siemens RBD IgG assay is a two-step sandwich indirect chemiluminescent assay with a manufacturer-reported 95.6% (95% CI: 92.2-97.8%) sensitivity and 99.9% (95% CI 99.6-99.9%) specificity for tests performed ≥21 days post positive RT-PCR test. An index value ≥1.0 is considered reactive and an index value of 150 is the upper limit of quantification. An index value of 1.0 corresponds to 21.8 binding antibody units (BAU)/mL according to the recently established World Health Organization (WHO) international standard^20^.

### Statistical Analysis

Authors MMR and JH led the statistical analyses. We described demographic data and laboratory values using proportions, mean (SD) or median, and 25^th^, 75^th^ percentile, as applicable. We first assessed the proportion of patients without a detectable IgG response over the follow-up period, overall and then stratified by RBD IgG antibody status prior to vaccination and vaccine type. We assumed presence of RBD IgG prior to vaccination indicated prior SARS-CoV-2 infection. Since patients on dialysis are tested monthly on or around the same date of each month, we reported data using discrete 30-day time windows post-vaccination, with the exception of the first time period in which we described data from the 14-30-day time period post-vaccination.

Next, among patients with a total RBD response, we computed median and 95% confidence intervals for RBD IgG index values using quantile regression with robust SEs to account for multiple observations per patient ^21^, as implemented in Stata *qreg* and *margins* commands. In this longitudinal data analysis, model parameters have a population-average interpretation^22^. We used quantile regression, and in particular, the median, to describe the data because it is invariant to data truncation and estimable in all of the analyses presented. We ascertained effect modification by separately adding to the quantile regression model the interaction between time window and RBD antibody status prior to vaccination, vaccine type, age, and diabetes status.

Finally, we used a nested case-control design with incidence density sampling to evaluate the association between post-vaccination infection and RBD IgG titers at two time points: the peak RBD IgG attained within the first 60 days post-vaccination, and the RBD IgG titer available in the most proximal time period prior to infection. The cohort comprised of fully-vaccinated patients. We set the time at which vaccination was completed to be time 0 (i.e., received two doses of the mRNA vaccines or a single dose of Ad26.COV2.S at least two weeks prior). We then defined cases as all patients in the cohort who had been identified with a COVID-19 diagnosis before September 14, 2021. We matched each case to five controls—i.e., all patients still alive, without infection, and receiving dialysis at a US Renal Care facility—by age (5-year category), sex, and calendar month of vaccination. Using conditional logistic regression further accounting for diabetes, we reported the odds of a COVID-19 case per five-unit change in index value, and for the discrete index value <10 (versus ≥10) and < 23 (versus ≥23). We selected the first cut-point based on data showing that index values ≥ 10 corresponded with pseudovirus neutralization titers^23^ > 1:500 and a positive predictive value of 100% for plaque reduction neutralization test_50_ > 1:80 in Siemens studies^24^. The latter cut point (23) corresponds to 506 binding antibody units/mL on the WHO international standard^20^, and was demonstrated by Feng *et al*. to correspond to 80% vaccine efficacy against symptomatic infection^12^.

## Results

Of the 4884 patients we selected to follow as of January 2021, 4791 (98%) were able to be followed (**Supplemental Figure 1**). Among these 4791, 2563 (54%) completed vaccination (**Table 1**). The remainder of the analyses evaluated antibody responses among this fully vaccinated cohort.

**Table 1.** Characteristics of patients on dialysis followed with monthly RBD antibody tests.

|  | **Vaccinated\***<br>**N=2563** | **Partially**<br>**vaccinated^**<br>**N=677** | **Unvaccinated**<br>**N=1551** | **Overall**<br>**N=4791** |
| --- | --- | --- | --- | --- |
| **Age (years)** | 64.7 (13.8) | 63.8 (13.4) | 61.0 (14.5) | 63.4 (14.1) |
| 18 to 44 | 202 (7.9) | 65 (9.6) | 232 (15.0) | 499 (10.4) |
| 45 to 64 | 1008 (39.3) | 258 (38.1) | 631 (40.7) | 1897 (39.6) |
| 65 to 79 | 980 (38.2) | 284 (42.0) | 529 (34.1) | 1793 (37.4) |
| ≥ 80 | 373 (14.6) | 70 (10.3) | 159 (10.3) | 602 (12.6) |
| **Women** | 1059 (41.3) | 288 (42.5) | 674 (43.5) | 2021 (42.2) |
| **Race and Ethnicity** |  |  |  |  |
| Hispanic | 357 (13.9) | 108 (16.0) | 172 (11.1) | 637 (13.3) |
| Non-Hispanic Black | 528 (20.6) | 177 (26.1) | 414 (26.7) | 1119 (23.4) |
| Non-Hispanic Other<sup>§</sup> | 431 (16.8) | 89 (13.2) | 165 (10.6) | 685 (14.3) |
| Non-Hispanic White | 776 (30.3) | 190 (28.1) | 457 (29.5) | 1423 (29.7) |
| Missing | 471 (18.4) | 113 (16.7) | 343 (22.1) | 927 (19.4) |
| **Region** |  |  |  |  |
| Northeast | 278 (10.9) | 57 (8.4) | 145 (9.4) | 480 (10.0) |
| South | 1045 (40.8) | 394 (58.2) | 933 (60.2) | 2372 (49.5) |
| Midwest | 281 (11.0) | 56 (8.3) | 136 (8.8) | 473 (10.0) |
| West | 959 (37.4) | 170 (25.1) | 337 (21.7) | 1466 (30.6) |
| **Diabetes** | 1514 (59.1) | 404 (59.7) | 852 (59.3) | 2770 (57.8) |
| Missing | - | - | 11 (0.7) | 11 (0.2) |
| **Vaccine Type** |  |  |  |  |
| mRNA1273 | 1259 (49.1) | 305 (45.1) | - | 1564 (32.6) |
| BNT162b2 | 1197 (46.7) | 361 (53.3) | - | 1558 (32.5) |
| Ad26.COV2.S | 107 (4.2) | - | - | 107 (2.2) |

\*Completed two doses of the mRNA vaccines or a single dose of the Ad2COVS vaccine at least 14 days prior to September 14, 2021. ^Received only a single dose of mRNA vaccine prior to September 14, 2021. <sup>§</sup>Persons self-reporting Asian, American Indian, Alaskan, and Pacific Islander heritages are captured as 'other.'

### Proportion with detectable post-vaccination RBD IgG

Among the vaccinated cohort, 20% had lost detectable RBD IgG response in the 5-6 months window post-vaccination (**Table 2**). A similar proportion of patients with and without prior exposure to SARS-CoV-2 infection lost detectable RBD IgG response post-vaccination (**Table 2**). A smaller proportion of patients receiving the mRNA1273 lost response in the 5-6 months window compared to BNT162b2 (11% versus 31%). For the Ad26.COV2.S vaccine, sufficient samples were available for the 4-5 month post-vaccination period, by which time 57% had lost a detectable RBD IgG response.

**Table 2.** Proportion of vaccinated patients on dialysis without RBD IgG^ over time.

|  | Days 14-30<br>N=1369 | Days 31-60<br>N=2304 | Days 61-90<br>N=2292 | Days 91-120<br>N=1861 | Days 121-150<br>N=1228 | Days 151-180<br>N=555 |
| --- | --- | --- | --- | --- | --- | --- |
| **RBD antibody status prior to vaccination** |  |  |  |  |  |  |
| Negative, n | 912 | 1601 | 1589 | 1346 | 974 | 503 |
| % without RBD IgG | 7.1 (5.6, 9.0) | 8.1 (6.9, 9.6) | 9.3 (8.0, 10.9) | 11.7 (10.1, 13.6) | 17.0 (14.8, 19.6) | 20.5 (17.2, 24.3) |
| Positive, n | 457 | 703 | 703 | 515 | 254 | 52 |
| % without RBD IgG | 5.7 (3.9, 8.3) | 6.0 (4.5, 8.0) | 6.7 (5.1, 8.8) | 8.7 (6.6, 11.6) | 14.2 (10.5, 19.2) | 17.3 (9.6, 31.4) |
| **Vaccine type** |  |  |  |  |  |  |
| mRNA1273, n | 605 | 1116 | 1105 | 1072 | 713 | 321 |
| % without RBD IgG | 2.3 (1.4, 3.9) | 3.0 (2.2, 4.2) | 4.5 (3.5, 5.9) | 5.5 (4.3, 7.1) | 8.6 (6.7, 10.9) | 10.9 (8.0, 14.9) |
| BNT162b2, n | 720 | 1033 | 1094 | 696 | 436 | 230 |
| % without RBD IgG | 6.3 (4.7, 8.3) | 8.0 (6.5, 9.9) | 9.3 (7.8, 11.2) | 13.6 (11.3, 16.5) | 22.0 (18.5, 26.3) | 31.7 (26.3, 38.4) |
| Ad26.COV2.S, n | 44 | 105 | 93 | 93 | 79 |  |
| % without RBD IgG | 72.7 (60.7, 87.2) | 51.4 (42.7, 61.9) | 46.2 (37.1, 57.6) | 52.7 (43.5, 63.9) | 57.0 (47.0, 69.0) |  |
| **RBD antibody status prior to vaccination** |  |  |  |  |  |  |
| **Negative** |  |  |  |  |  |  |
| mRNA1273, n | 409 | 823 | 786 | 773 | 568 | 288 |
| % without RBD IgG | 2.7 (1.5, 4.8) | 3.5 (2.5, 5.0) | 5.5 (4.1, 7.3) | 6.3 (4.8, 8.3) | 9.3 (7.2, 12.1) | 10.8 (7.7, 15.0) |
| BNT162b2, n | 477 | 711 | 745 | 512 | 355 | 213 |
| % without RBD IgG | 7.1 (5.2, 9.9) | 9.4 (7.5, 11.8) | 10.6 (8.6, 13.1) | 15.0 (12.2, 18.5) | 23.7 (19.6, 28.5) | 32.9 (27.1, 39.8) |
| Ad26.COV2.S, n | 26 | 67 | 58 | 61 | 51 | - |
| % without RBD IgG | 77.0 (62.3, 94.9) | 50.7 (40.1, 64.2) | 44.8 (33.7, 59.6) | 52.5 (41.3, 66.6) | 56.9 (44.8, 72.2) | - |
| **Positive** |  |  |  |  |  |  |
| mRNA1273, n | 196 | 343 | 319 | 299 | 145 | 33 |
| % without RBD IgG | 1.5 (0.5, 4.7) | 1.7 (0.8, 3.9) | 2.2 (1.1, 4.6) | 3.3 (1.8, 6.2) | 5.5 (2.8, 10.8) | 12.1 (4.8, 30.4) |
| BNT162b2, n | 243 | 322 | 349 | 184 | 81 | 17 |
| % without RBD IgG | 4.5 (2.5, 8.1) | 5.0 (3.1, 8.0) | 6.6 (4.4, 9.8) | 9.8 (6.3, 15.2) | 14.8 (8.8, 25.0) | 17.6 (6.3, 49.3) |
| Ad26.COV2.S, n | 18 | 38 | 35 | 32 | 28 | - |
| % without RBD IgG | 66.7 (48.1, 92.4) | 52.6 (38.9, 71.2) | 48.6 (34.5, 68.3) | 53.1 (38.4, 73.6) | 57.1 (41.5, 78.8) | - |
| **Overall % without RBD** | **6.6 (5.5, 8.1)** | **7.5 (6.5, 8.6)** | **8.5 (7.4, 9.7)** | **10.9 (9.6, 12.4)** | **16.4 (14.5, 18.7)** | **20.2 (17.1, 23.8)** |

^Includes lack of seroconversion on a total RBD assay or with RBD IgG index value below assay limit (<1). Data in parentheses represent 95% CI of the proportion without RBD IgG. Missing data are for time points with insufficient data.

### Quantified post-vaccination RBD IgG values

RBD IgG index values declined over time among patients with a detectable total RBD antibody response. By 5-6 months post-vaccination, the median index values for the overall cohort were 3.7 (95% CI: 3.1, 4.3) compared with a median peak index value of 92.7 (95% CI: 77.8, 107.5) in the 14-30 days after completing vaccination (**Figure 1a**). The overall trajectory of the response differed by SARS-CoV-2 infection status prior to vaccination, vaccine type, and age groups (p-value for interaction for each subgroup <0.001). The peak response was higher among patients with evidence of prior SARS-CoV-2 infection but by end of follow-up there was no difference in index values among patients with and without prior SARS-CoV-2 infection (**Figure 1a**, p-value for difference in end of follow-up median index values=0.230). Patients receiving the mRNA1273 vaccine had higher median RBD IgG index values throughout follow-up (**Figure 1b**, p-value for difference in end of follow-up <0.001). Older patients (≥80 years old) had lower peak index values and slightly lower median index values at the end of follow-up (**Figure 1c;** p-value for difference in end of follow-up =0.0288). There was no difference in peak or median index values over time by diabetes status (**Figure 1d**).

**Figure 1:**
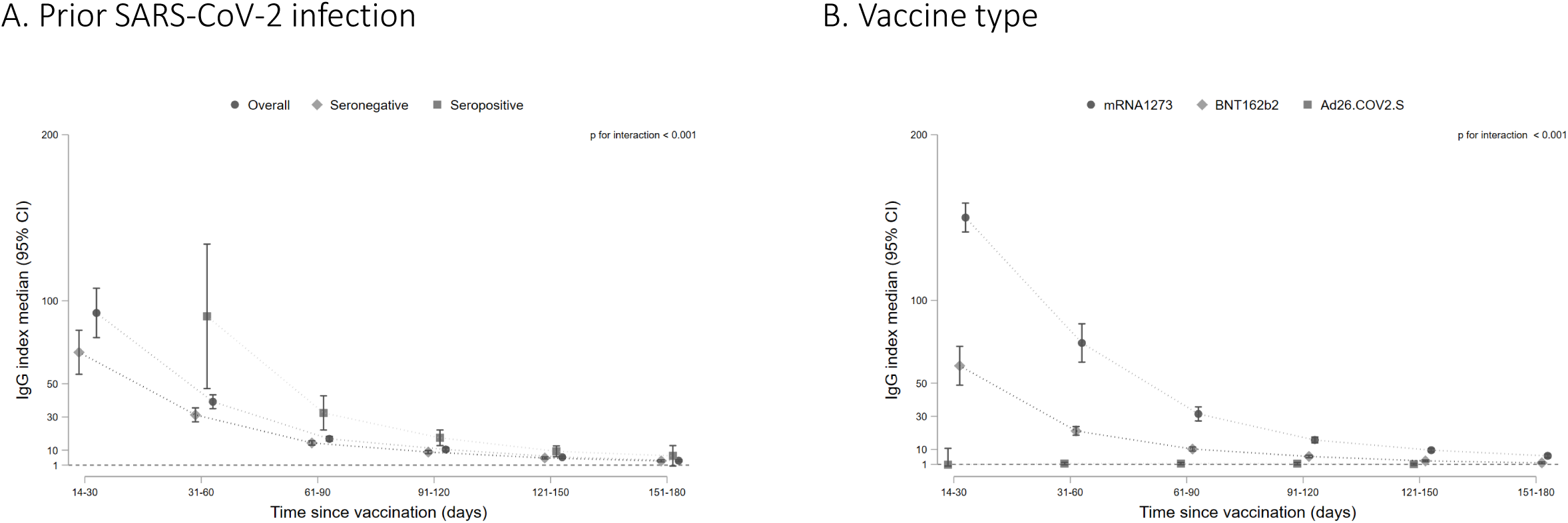
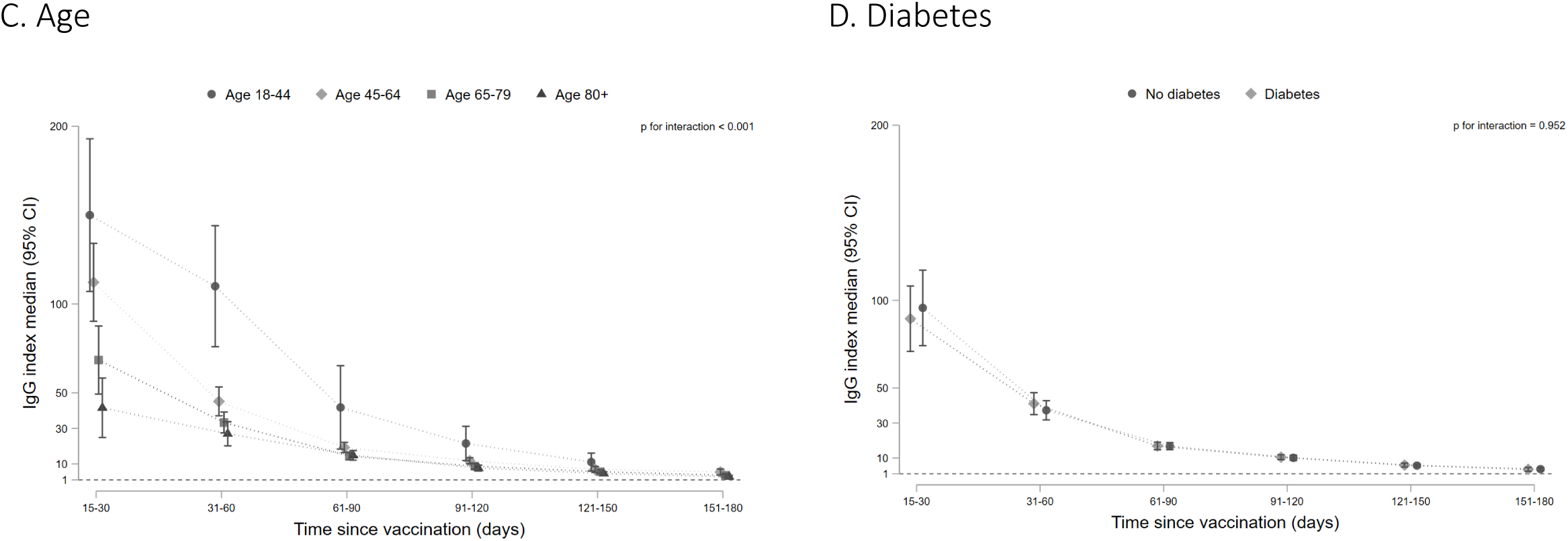
RBD IgG index values over time among patients on dialysis in the overall cohort and by prior SARS-CoV-2 infection (A), by vaccine type (B), by age group (C), and diabetes status (D). Among patients who seroconverted on the total RBD Ig assay, median RBD IgG index values are graphed by time since vaccination, with error bars representing 95% CI for the median value. A missing time point indicates insufficient data for the subgroup at that time point. An index value of 1 corresponds to 21.8 BAU/mL on the WHO Standard. Index values < 1 indicate a ‘negative’ result on the assay. P values test for interaction by subgroup and significant p values indicate that the trajectory of the response differed by the subgroup depicted.

### Association of post-vaccination RBD IgG values with breakthrough infection

As of September 14, 2021, there were 56 cases of breakthrough COVID-19 (median time from vaccination to infection: 110 [25^th^-75^th^ percentile 85-143] days). Of these, 25 (45%) were hospitalized with admission or discharge diagnoses indicating COVID-19 and an additional 15 (27%) had documented symptoms (fever, cough, and chills being the most common). The matching procedure achieved balance in distribution of age, sex, and calendar month of vaccination among cases and controls (**Supplemental Table 2**). Cases had lower peak and pre-breakthrough RBD IgG index values compared with controls (**Figure 2a, b**). Low pre-breakthrough index values were associated with breakthrough infection among cases (**Table 3**). Similar associations were observed with peak IgG values. All cases had pre-breakthrough index values < 36 (equivalent to 785 BAU/mL). Based on the conditional logistic regression model, the predicted probability of being a case at index values > 36 was <5% (**Supplemental Figure 2**).

**Figure 2:**
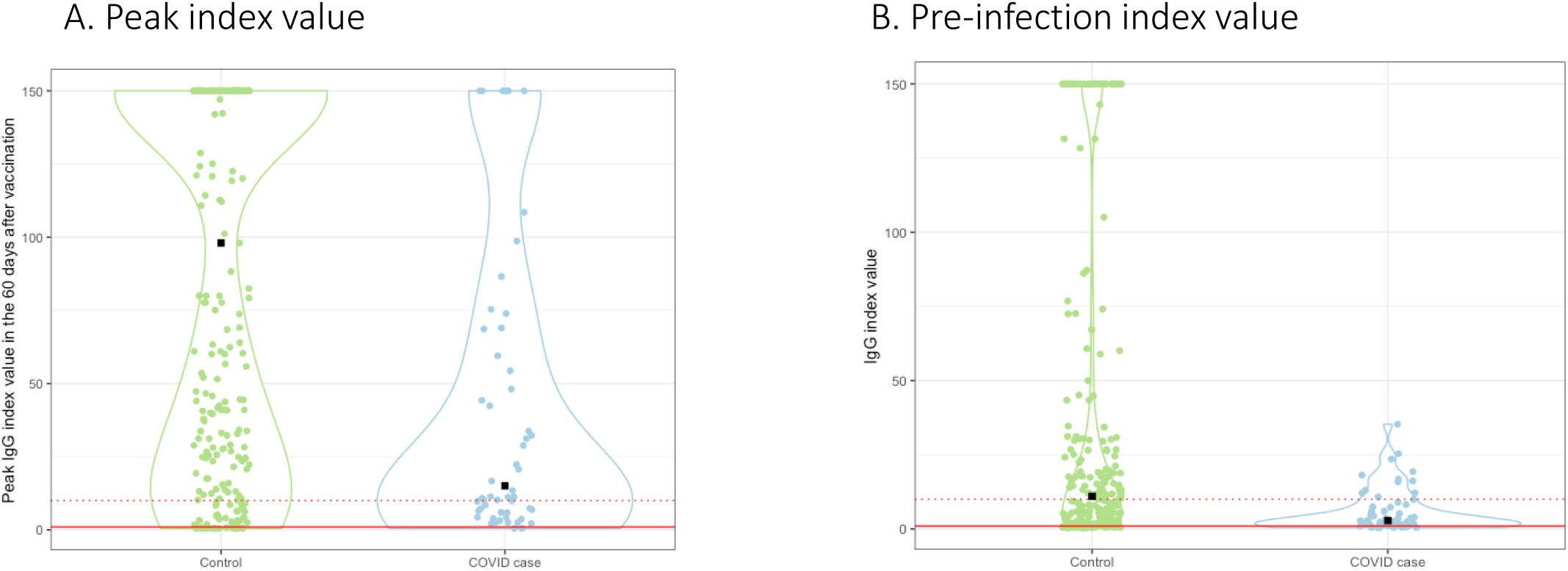
RBD IgG index values among cases versus controls. Violin plots represent peak RBD IgG values obtained within 60 days prior to vaccination (A) and the IgG values obtained in immediate period preceding infection are graphed (B) by case versus control status. Median time between pre-breakthrough IgG values and COVID-19 diagnosis was 21 days (25^th^, 75^th^ percentile: 14-28 days); the corresponding time for controls was 21 days (25^th^, 75^th^ percentile 12, 27 days). Median peak RBD IgG values were 98.0 (22.3, 150) versus 15.1 (6.3, 71.5), and pre-breakthrough values were 11.0 (2.3, 47.8) versus 2.8 (1.2, 8.6) for controls versus cases respectively. All cases had index values < 36.

**Table 3.** Association of peak and pre-breakthrough RBD IgG index values with post-vaccination in infection in patients on dialysis^.

| **RBD IgG index value** | **Odds ratio (95% CI)** | **p-value** |
| --- | --- | --- |
| **Peak \*** |  |  |
| Below 10 (vs. $\geq 10$) | 2.68 (1.45, 4.97) | 0.002 |
| Below 23 (vs. $\geq 23$) | 3.70 (2.02, 6.79) | <0.001 |
| **Pre-breakthrough \*** |  |  |
| Below 10 (vs. $\geq 10$) | 5.02 (2.38, 10.58) | <0.001 |
| Below 23 (vs. $\geq 23$) | 9.80 (2.94, 32.76) | <0.001 |

<sup>^</sup>Case control analysis in which cases were matched 1:5 to controls by age, sex, and calendar month of vaccination. \*Associations determined from separate models which adjust for diabetes.

## Discussion

In our diverse, national cohort of patients receiving dialysis, one in five have lost a detectable RBD antibody response within a 6-month period post-vaccination. Low levels of circulating RBD antibody as measured using a high-throughput assay were associated with risk for breakthrough infection. Although peak median antibody values were higher among patients with prior-to-vaccine SARS-CoV-2 infection, the proportion without a detectable response and median antibody values were similar among the patients with and without prior SARS-CoV-2 infection by the end of follow-up. Vaccine type was associated with the strength of the RBD antibody response throughout follow-up, with mRNA1273 conferring the highest median RBD index values and the lowest proportion of patients without a detectable response.

Large national studies^25,26^ demonstrating decreasing vaccine effectiveness in parallel with a decline in post-vaccination antibody response^27^ provide increasing data to support that post-vaccination antibody response could be useful correlate of protection. Experts have further posited that antibody correlates of protection against breakthrough COVID-19 infection could offer surrogate outcomes for vaccine development, assess population-level immunity, and inform immunization schedules^20,28^. However current data—which emphasize neutralizing antibodies^12,29,30^, or draw from clinical trials^11,12^ or health care worker cohorts^13^ do not have broad applicability. Furthermore most studies evaluate peak or early (<60-day) response to vaccination^11,12^; this measurement will no longer be available for vaccinated populations.

In our cohort, we were able to implement an unbiased monthly serologic testing strategy to study post-vaccination response in a geographically diverse population with sizeable proportions of racial and ethnic minority groups, and of patients with chronic illnesses (e.g., heart failure or diabetes). Using these real-world data from a time at which the delta variant of SARS-CoV-2 was also in wide circulation in the US, we found a clinically meaningful indication that antibody values measured using an accessible assay and at time points remote from vaccination are strongly associated with risk for breakthrough infection. This brings us closer to defining a ‘persisting antibody’ threshold^31^ for immunity. The relative importance of such a threshold may be greater for high-risk or immunocompromised groups compared with otherwise healthy persons since many components of their immune response may be impaired^32^. This is evident in our data since 40% of patients with breakthrough infection were hospitalized.

Although all of our cases had RBD IgG index values < 36, a substantial number of controls also had low antibody response measured contemporaneously. This is concordant with other data^11,13^, and may imply that sensitivity of any single selected persisting antibody threshold may be low. However, it also possible that many more of our control patients with low circulating antibodies would have developed COVID-19 absent other mitigation strategies such as masks or social distancing. Given that up to one-quarter of patients on dialysis hospitalized with COVID-19 died^33^, and similar or higher mortality rates are described among immunocompromised groups with COVID-19 infection, it is critical that we identify those at high risk who need heightened protection. That antibody thresholds may lack sensitivity is counterbalanced by the relatively low risk associated with enhanced mitigation for those classified as ‘at risk’. Research could also explore whether serologic testing improves uptake of the newer recommendations to add doses among select groups.

Several studies have reported a stronger initial antibody response with mRNA1273 compared with BNT162b2, putatively due to the higher mRNA dose in mRNA1273 formulation^32,34^. We found that mRNA1273 vaccinated persons maintained slightly higher index values than BNT162b2 throughout the 6-month follow-up. A single dose of Ad26.CoV2.S did not yield a detectable antibody response in more than half of patients. Ad26.CoV2.S manufacturers have recently submitted data that demonstrate the improved efficacy of this vaccination with two doses given two months apart^35^. Finally, although healthy persons who experience SARS-CoV-2 infection prior to vaccination seem to mount peak antibody responses more than two-fold higher than those who were not infected^36^, we find that antibodies among patients on dialysis wane over time irrespective of prior SARS-CoV-2 infection and are equivalently low six months after vaccination. Patients with prior SARS-CoV-2 infection also experienced breakthrough infection in our cohort.

The limitations of our work include the reliance on RBD as the single measure of antibody response, and use of the electronic health records to detect cases. Although pragmatic and likely detecting clinically meaningful events, this strategy misses asymptomatic cases and those with minor symptoms. Vaccine types were not randomly allocated and the number of patients who received Ad26.CoV2.S was low; therefore, comparisons of response by vaccine type should be interpreted with caution. The number of breakthrough infections was relatively small, but was proportionately higher than in healthy cohorts, and in absolute numbers equivalent to or higher than those described from clinical trials^11,12^. Furthermore, the observed associations despite the modest sample size and number of events highlights the relevance of our findings. Finally recommended immunization schedules are changing, with an additional (third) dose of BNT162b2 now added for select groups in the US, and our study does not evaluate response to these third doses.

In summary, among patients receiving dialysis, the humoral response to SARS-CoV-2 vaccination wanes and is associated with risk for breakthrough infection. Serologic testing using commercially available high-throughput assays could inform vaccination and enhanced mitigation strategies in immunocompromised and other high-risk populations. Even as new vaccine platforms and immunization schedules are employed, further research investigating the associations between antibody response and risk for infection may guide management, particularly for immunocompromised hosts.

## Data Availability

Data may be available upon review and approval of analysis request by all co-authors. Data may require data sharing agreements.

## Disclosures

LC, PH, CM, RK and PB are employed by Ascend Clinical Laboratories. MD and GB are employed by US Renal Care. GC is on the Board of Satellite Healthcare, a not for profit dialysis organization.

## Funding Support

Ascend Clinical Laboratory supported the remainder plasma testing for SARS-CoV2 antibodies.

Dr. Anand was supported by R01DK127138. Dr. Garcia is supported by the Leeds Scholarship Fund. Dr Chertow was supported by K24DK085446.

## Notes

### Author Declarations

The study received ethics approval from Stanford University. Stanford University investigators received anonymized data, and the Institutional Review Board waived requirement for consent.

