## supplemental tables 1, 2, figure 1, 2 for "SARS-CoV-2 vaccine antibody response and breakthrough infection in dialysis"

**Table of Contents**

**Supplemental Methods** Sample size of cohort selected for monthly antibody monitoring	p3

**Supplemental Figure 1.** Study flowchart of participants	p5

**Supplemental Table 1** Distributions of age, sex and diabetes mellitus status in the cases, controls and overall.	p6

**Supplemental Figure 2:** Predicted probabilities for case versus control status for observed RBD IgG index values	p7

**Supplemental Methods: Sample size of cohort selected for monthly antibody monitoring**

Our study was conducted in partnership with the dialysis network US Renal Care and Ascend Clinical Laboratory. Ascend Clinical tested remainder plasma of patients for SARS-CoV-2 antibody, and anonymized all patient demographic, comorbidity, and laboratory data prior to transfer to Stanford University. We selected 4,348 patients on follow from the 17,390 patients on dialysis without prior evidence of SARS-CoV-2 infection (as of January 2021) in US Renal Care network. To estimate sample size, we used previously published data on hepatitis B vaccination non-response, as available by age strata from Bruguera et al.^1^, who evaluated immune response in 270 patients. In this study, the rate of non-response among persons age 20-40, 40-60, and >60 years was 7%, 13%, and 35% respectively. Correspondingly, our estimates of non-response among persons age 18-44, 45-64, ≥65 years was 5%, 15%, and 30% respectively. Estimating these proportions of non-response with an absolute precision of 2%, and oversampling by 15%, resulted in a sample size estimate of 4222 (see Table).

Population and subpopulation sizes by age and number of patients required to obtain a prevalence estimate with the specified absolute precision assuming and the specified proportion of non-response to the vaccine.

| **Age group** | **Proportion of non-response to vaccine** | **Absolute precision** | **USRDS Population count** | **US Renal Care Population size** | **Sample size required** | **Over sample (15%)** |
| --- | --- | --- | --- | --- | --- | --- |
| 18 to 44 | 5% | 2% | 60,540 | 2,871 | 453 | 521 |
| 45 to 64 | 15% | 2% | 207,022 | 10,605 | 1,218 | 1,401 |
| ≥ 65 | 30% | 2% | 231,588 | 12,777 | 2,000 | 2,300 |
| Total |  |  | 499,150 | 26,253 | 3,671 | 4,222 |

We used systematic sampling with fractional intervals. In systematic sampling the patients are selected from the list using a fixed selection interval, calculated by dividing the total number of patients in the list by the desired number (i.e., 17390/4222 = 4.1). We thus randomly selected one number between 1 and 4 and then selected every 4th patient in the sampling frame sorted by zip code, age, sex and race. This resulted in a sample size of 4344 patients on dialysis. In addition, since August 2020, we have followed 6551 patients among whom a subset seroconverted prior to vaccination (i.e., developed evidence of natural infection). All 540 US Renal Care patients seropositive as of January 2021 were also selected to be followed.

This resulted in a final sample of 4884 patients in whom we initiated monthly antibody testing, 4791 were able to be followed starting February 2021, and 2563 were vaccinated with at least 14 days of available data as of September 14 2021 (see Supplemental Figure 1 for final vaccinated cohort).

**Supplemental Figure 1: Flowchart of study participants**

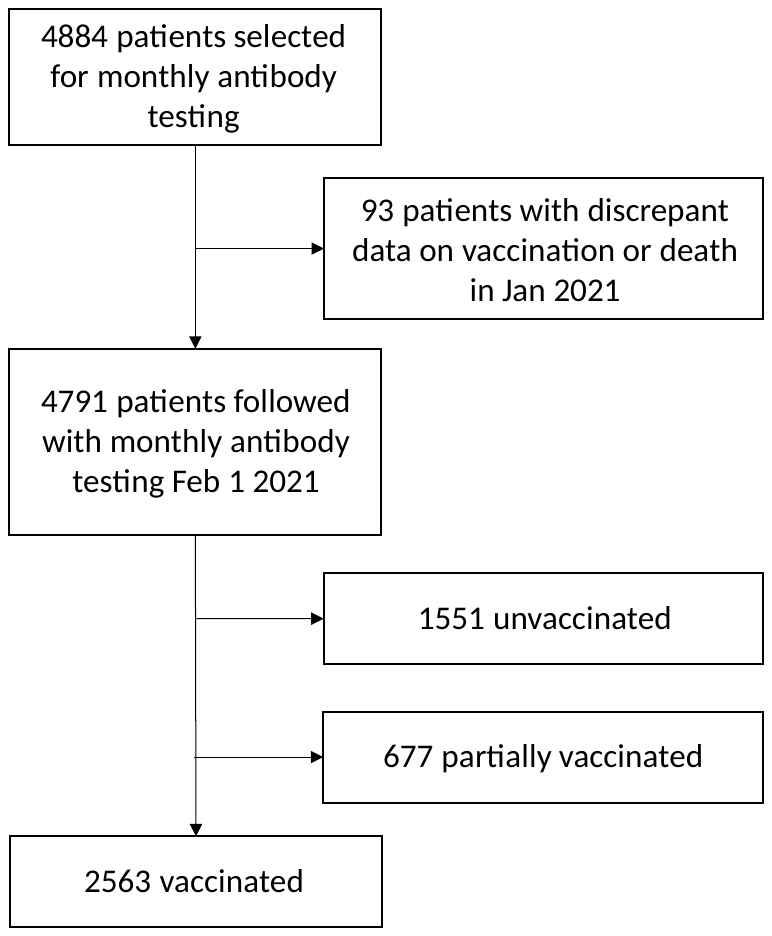

**Supplemental Table 1.** Distributions of age, sex and diabetes mellitus status in the cases, controls and overall.

| Patient characteristics | Cases  N=56 | Controls  N=280 |
| --- | --- | --- |
| **Age, mean (SD)** | 66.2 (13.3) | 66.0 (13.6) |
| **Age (5 year frequencies, %)** |  |  |
| 35 | 5 | 5 |
| 45 | 29 | 29 |
| 60 | 5 | 5 |
| 65 | 21 | 21 |
| 70 | 14 | 14 |
| 75 | 7 | 7 |
| 80 | 7 | 7 |
| 85 | 7 | 7 |
| 90 | 4 | 4 |
| **Women , %** | 38 | 38 |
| **Month vaccination completed, %** |  |  |
| February | 16 | 16 |
| March | 14 | 14 |
| April | 23 | 23 |
| May | 46 | 46 |
| **Diabetes Mellitus, %** | 71 | 65 |
| **RBD antibody seropositive prior to vaccination, %** | 20 | 35 |

**Supplemental Figure 2: Predicted probabilities for case versus control status for observed RBD IgG index values**

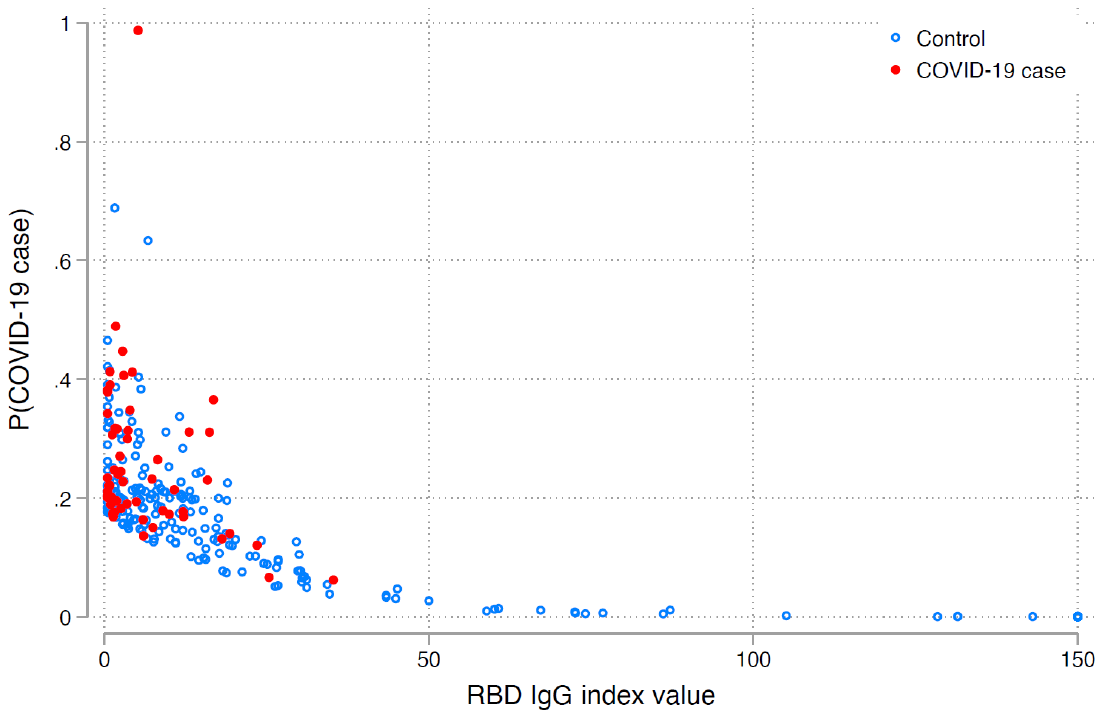

In the conditional logistic model evaluating 56 matched sets of cases and controls, the probability of being a case at index values > 36 was < 5%; it was 0% for index values > 50.
